# Modelling the impact of vaccination and sexual behavior change on reported cases of mpox in Washington D.C

**DOI:** 10.1101/2023.02.10.23285772

**Authors:** Patrick A. Clay, Jason M. Asher, Neal Carnes, Casey E. Copen, Kevin P. Delaney, Daniel C. Payne, Emily D. Pollock, Jonathan Mermin, Yoshinori Nakazawa, William Still, Anil T. Mangla, Ian H. Spicknall

## Abstract

**Background:** The 2022 mpox outbreak infected over 30,000 people in the United States. Infections were commonly associated with sexual contact between men. Interventions included vaccination and reductions in sexual partnerships. We estimated the averted infections attributable to each intervention using mathematical modeling.

**Methods:** We fit a dynamic network transmission model to mpox cases reported by the District of Columbia through January 2023. We incorporated vaccine administration data and reported reductions in sexual partnerships among gay, bisexual, or other men who have sex with men (MSM). Model output consisted of predicted cases over time with or without vaccination and/or behavior change.

**Results:** We estimated initial case reductions were due to behavior change. Vaccination alone averted 64% [IQR:57%-72%] and behavior change alone averted 21% [IQR:11%-29%] of cases. Vaccination and behavior change together averted 80% [IQR:74%-85%] of cases. In the absence of vaccination, behavior change reduced cumulative cases but also prolonged the outbreak.

**Conclusions:** Initial case declines were likely caused by behavior change, but vaccination averted more cases overall. Overall, this indicates that encouraging individuals to protect themselves was vital in the early outbreak, but that combination with a robust vaccination program was ultimately required for control.

## Introduction

Since May 2022, over 30,000 cases of mpox have been reported to the Centers for Disease Control and Prevention (CDC). The majority of infections have been among gay, bisexual, and other men who have sex with men (MSM) and have been associated with sexual contact^1,2^. Incidence decreased by >80% from a peak of 440 daily cases in mid-August to 70 in mid-October and fewer than 4 daily cases nationally as of January 2023^3^. In response to the mpox outbreak, CDC and other public health authorities recommended sexual behavioral change and after the JYNNEOS™ (Modified Vaccinia Ankara vaccine, Bavarian Nordic) mpox vaccine became increasingly available, preventive vaccination. In early June, CDC developed messaging for individuals seeking to reduce their chances of acquiring mpox (updated on August 5)^4^. CDC additionally worked with partner organizations, media, and digital apps to communicate this information to MSM, the most affected population^2^. Later surveys reported that 50% of MSM who engaged in one-time sexual encounters had reduced their frequency of these encounters since the beginning of the mpox outbreak^5^. In addition to behavioral interventions, JYNNEOS™ vaccine became available from the Strategic National Stockpile for post-exposure prophylaxis shortly after the first confirmed case of mpox on May 17^th^^6,7^, and for pre-exposure prophylaxis for impacted communities in late June.

Understanding the relative effects of sexual behavior change and vaccine-induced immunity on mpox cases can inform public health efforts for this and future outbreaks. A previous study suggested that mpox outbreaks in the United States and other countries could have ended due to infection-driven herd immunity alone, but did not incorporate the potential role of behavioral change or vaccination in outbreak trajectory^8^. We used data from Washington, DC, and a dynamic network model of sexually transmitted infection (STI) transmission to estimate the relative effects of behavior change and vaccination on the outbreak, and the theoretical impact if vaccines had been more fully available earlier in the outbreak.

## Methods

### Model Overview

We adapted a previously published dynamic network model of STI transmission in MSM^9,10^ to mpox transmission^11^ and added behavioral change and vaccination. The contact structure of the network is based on surveys of MSM recruited through websites^9^ or at Lesbian, Gay, Bisexual, Transgender, Queer, and other gender and sexual minorities (LGBTQ+) serving establishments in Atlanta, Georgia^12^. Participants were asked whether they had a “main” partner (defined as an enduring sexual partner who took priority over others), how many “casual” partners they had (defined as enduring sexual partners other than “main” partners), and how many one-time sexual contacts they had had in the past three months. Based on survey results, individuals in the model have a set probability of having 0 or 1 main partners with a mean relationship duration of 477 days, and 0-2 casual partners with a mean relationship duration of 166 days. Additionally, individuals in the model are assigned to 1 of 6 sexual activity groups, depending on their probability per day of engaging in one-time sexual partnerships, with activity group 1 representing the lowest sexual activity level (never engaging in one-time partnerships) and activity group 6 representing the highest sexual activity level (29% chance of engaging in one-time partnership per day). Individuals have a probability of forming one-time partnerships during each model timestep based on their activity group and their number of main and casual partnerships (Table S1). Individuals choose partners based on age and preferred sexual position during intercourse (purely insertive, purely receptive, or versatile).

Individuals in the model can exist in the following states: susceptible to infection (*S*), infected but pre-symptomatic and non-infectious (*E*), pre-symptomatic and infectious (*P*), symptomatic and infectious (*I*), recovered and resistant (*R*), and vaccinated with one (*V*1) or 2 (*V*2) doses of JYNNEOS. We assume that symptom onset lowers the probability of an individual engaging in one-time sexual contact, modeled as individuals temporarily moving to one lower sexual activity level. We further assume that symptoms reduce the probability of sexual contact with main and casual partners by half. At each timestep, the probability that an infectious individual will infect a susceptible partner is the product of the probability of sexual contact per timestep in each relationship type and the probability of infection per sexual contact (*μ*). Upon infection, an individual enters the *E* state, after which they have a probability per timestep of entering the *P* and then *I* and *R* states. Individuals have a probability of seeking medical attention upon symptom onset (*ρ*); we assume those who seek care become aware of their infectious status, and so do not engage in one-time or casual sexual partnerships until they recover. We assume that individuals who learn their infection status also decrease contact with their main partners such that they have a 10% chance of infecting their main partner over the duration of the infection, reflecting prior estimates of household transmission^13^. Infected individuals who seek medical care are reported as diagnosed cases, hereafter referred to as *cases*.

Our model represents an outbreak in a single well-connected population, and thus best represents outbreak dynamics within a single city, rather than at the national scale. We parameterized the model to represent the mpox outbreak in Washington D.C., which reported its final case on Nov. 22, 2022. Thus, we set the population size to be 37,400, roughly equal to the population of MSM in Washington D.C.^14^. We additionally ran our analysis for a population size of 20,000, as not all MSM may be in the relevant sexual mixing pool (Fig. S4), and found that our qualitative results do not change. The model begins with 5 newly infectious individuals randomly selected from the 8,976 individuals in activity groups 5 and 6 on May 21^st^. This produced a median of 5 cumulative cases in the model on June 6^th^, matching D.C. data. The length of time between symptom onset and medical attention seeking is derived from the average time between symptom onset and orthopoxvirus test reported by Washington D.C. (Fig. S3).

### Vaccination

As input for our model, we took first and second doses administered in Washington D.C. through December 10, at which time vaccination rates had dropped from an approximate maximum of 4,500 doses given per week to fewer than 100 doses given per week. Pre-exposure vaccination for MSM began the week of June 26^th^ and peaked the week of July 31^st^ for first doses, and peaked the week of August 28^th^ for second doses (Table S2). As vaccines were originally offered primarily to MSM with multiple recent sexual partners^15^, we model vaccine distribution as limited to only the top two sexual activity groups for the first four weeks of vaccination, as limited to the top four sexual activity groups for the next four weeks of vaccination, and as limited to any individuals with a non-zero probability of engaging in one-time sexual contacts for the rest of the time period. In our model, we vaccinate susceptible (*S*) and pre-symptomatic (*E, P*) individuals. Individuals who receive the vaccine while in the *E* or *P* class are not prevented from becoming infectious. For second dose vaccination, we randomly draw individuals from the pool of those who have received their first dose at least four weeks in the past. We assume that the vaccine has no efficacy for two weeks after the first dose, at which time efficacy jumps to 37%, and that efficacy increases again to 69% two weeks after the second dose based on U.S. data^16^, though we run a sensitivity analysis based on an 85% first dose efficacy based on data from Israel^17^ (Fig. S8). In the case of breakthrough infections, individuals enter the pre-symptomatic (*E*) class and prior vaccination has no further effect on subsequent contagiousness or pathogenicity. We assume that some reported vaccinations are administered to non-msm or non-resident commuters rather than the modeled population. Thus, we fit the total proportion of reported vaccines given to the modeled population to case data, as described below.

### Behavior Change

Individuals may change sexual behavior in response to perceived risk level^18^. In our model, we assume that MSM reduced their probability of one-time sexual contacts per day by a time-varying relative percentage. For tractability, we assume all individuals reduce their probability of one-time contact by the same relative amount, resulting in the largest reduction in sexual partner numbers among high-activity individuals. To measure risk perception, we identified LGBTQ+ focused communities on the social media discussion site Reddit (“subreddits”) that contained multiple discussions about mpox. We used the number of posts and comments in these subreddits containing the terms “monkeypox”, “mpox”, or “mpx” over time (hereafter “Reddit activity”) as a proxy for risk perception (appendix). The relative reduction in sexual activity per day in our model was set equal to relative Reddit activity. We fit maximum reductions in sexual activity to case data (described below).

### Modelling Process

We fit five parameters to D.C. case data: the probability of transmission per contact (*μ*), the average length of the pre-symptomatic infectious period (*l*), the number of individuals infected with mpox virus at Capital Pride Parade/Festival, occurring on June 11-12 (*ε*), the maximum percent reduction in probability of one-time sexual contact per day in response to the mpox outbreak (*ω*), and the proportion of reported vaccine doses given to the modeled population (*θ*) (see appendix for details). We additionally fit our model to case data assuming no interventions to test the likelihood that the outbreak ended due to infection-driven herd immunity alone.

We then estimated what proportion of cases have been averted by either behavior change or vaccination. We ran the model with no vaccination and no behavior change. This represents a scenario in which no interventions had taken place. We then compare the cumulative cases in this model until May 21^st^ 2023 with cases in models where we include behavior change, include vaccination, or include both interventions. Differences in cumulative cases between intervention scenarios and the no intervention scenario represent cases averted by these interventions. We additionally estimated cumulative cases if vaccinations had been rolled out 28 days earlier or later, and if available vaccine doses were increased or decreased by 50% (behavior change was included in this sensitivity analysis). For each intervention scenario, we calculated median and interquartile ranges for cumulative case data over time for 120 model runs.

## Results

Fitting the model with vaccination and behavior change to cumulative case data from Washington D.C., we find that an 87.5% probability of transmission per sex act (*μ*), a pre-symptomatic infectious period (*l*) lasting 4 days, 40 individuals infected at D.C. Pride (*ω*), a 40% maximum reduction in probability of one-time sexual contacts in response to the mpox outbreak (*ε*), and 100% of reported vaccines being administered to the modeled population (*θ*) generates the best fit between model output and empirical data (Figure 1A). This indicates that mpox virus is highly transmissible, and that MSM substantially adjusted their behavior to reduce onward transmission of mpox virus. While it is implausible that 100% of reported vaccine doses were administered to resident MSM in D.C. rather than healthcare workers and non-resident MSM, a fit *θ* value of 100% indicates that a negligible amount of reported vaccine doses were administered to non-modelled populations (see Fig. S5 for proportion population vaccinated over time). We found a relatively low likelihood that the outbreak ended due to infection-driven herd immunity alone (Fig. 1, A vs. B).

**Figure 1:**
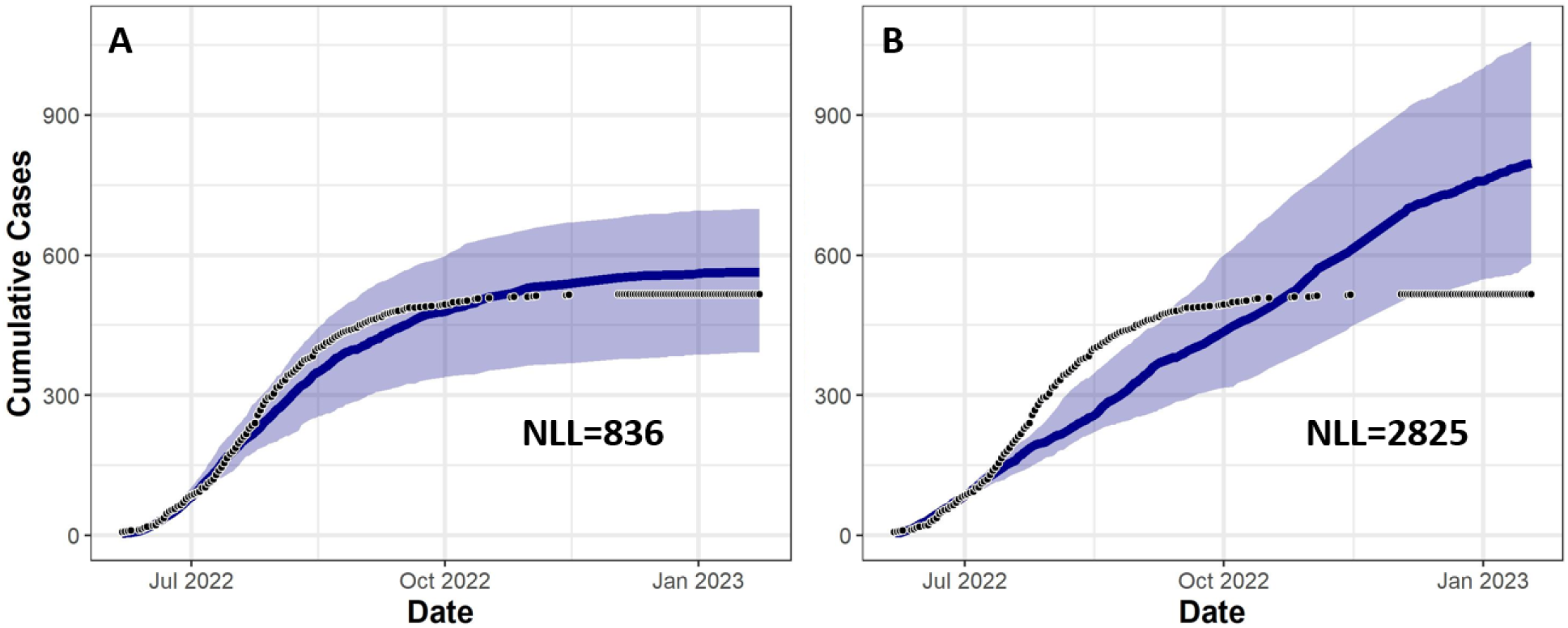
Model output fit to cumulative cases from Washington D.C. Y-axis shows cumulative cases, over date on the X-axis. Black dots represent cumulative case reports from Washington D.C., while the blue line represents median cumulative cases over 120 model runs for the best fitting parameter set, and the blue band represents the interquartile range, when (A) we fit all parameters, including effectiveness of interventions, and (B) we assume no intervention effectiveness. NLL indicates negative log likelihood, with lower values indicating a higher likelihood.

### Cumulative Cases

We estimated that initial reductions in mpox cases were due to behavioral change. The trajectory of the “behavior change only” model matches the “vaccine and behavior” model until September for cumulative cases, indicating that until this point, reductions in case reports were due primarily to behavior change (Figure 2A). The “vaccine only” scenario, on the other hand, closely matched the “no intervention” scenario until September for cumulative cases, indicating that behavior change altered the epidemic trajectory before vaccination. Behavior change has an earlier impact on cases than vaccination for two reasons. First, we find that Reddit activity, and thus behavior change, began when cases first appeared in D.C. (Fig. S1), whereas vaccine supply did not reach more than 400 doses per week until July. Second, reductions in sexual partnerships immediately reduce transmission, whereas we assume no protection from vaccination until 14 days after the first dose.

**Figure 2:**
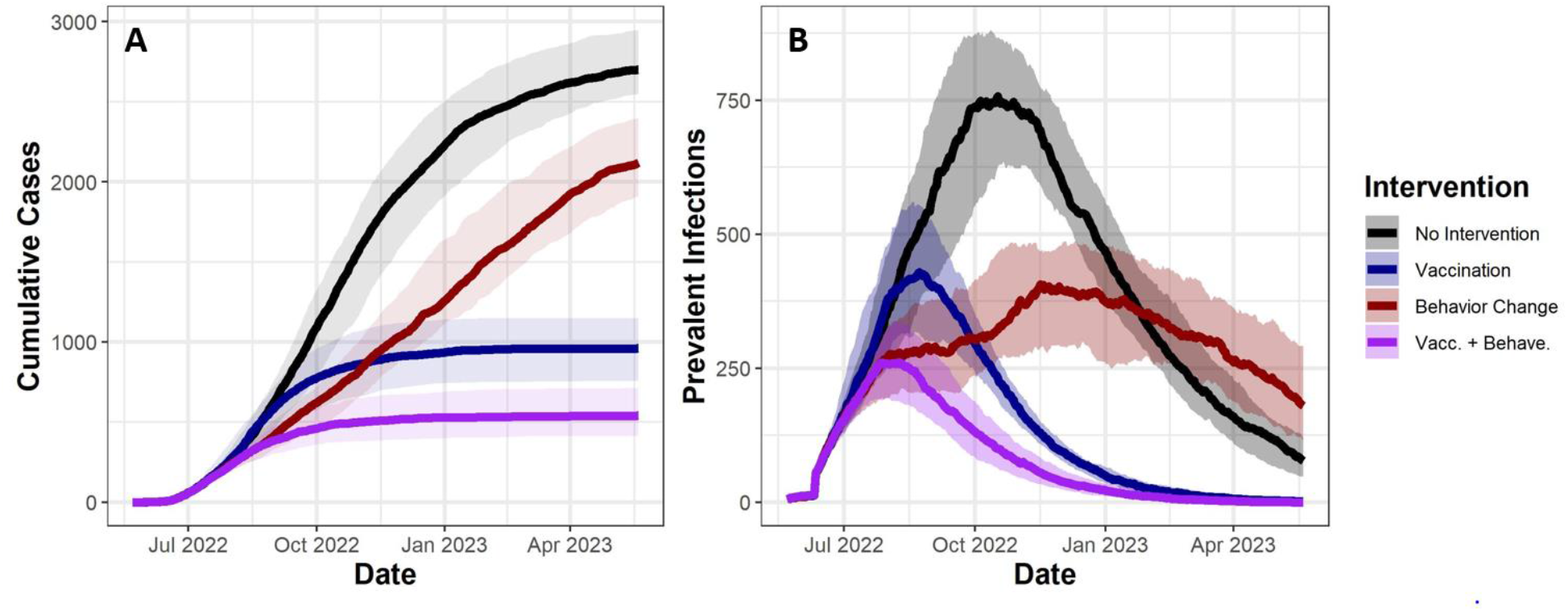
Vaccination and behavior change both reduce estimated cumulative cases and prevalent infections. (A) Y-axis shows model estimates of cumulative cases (i.e. individuals who are diagnosed with mpox), over time on the X-axis from May 21^st^ 2022 to May 21^st^ 2023. (B) Y-axis shows prevalent infections, over time on the X-axis. Solid lines indicate median values from 120 simulations, while transparent bands represent interquartile ranges. Colors indicate intervention combinations.

We estimated that one year into the outbreak, vaccination will have prevented more cases than behavior change. We estimated that by May 21^st^ 2023, we would have 2,700 [IQR 2,550 – 2,950] cumulative case reports in the absence of both interventions. The model estimated that behavior change alone would have prevented 21% [IQR 11% - 29%] of cases, vaccination alone would have prevented 64% [IQR 57% - 72%] of cases, and both interventions together will have prevented 80% [IQR 74% - 85%] (Fig. 2A).

In our model, behavior change helped vaccination avert more infections by allowing more individuals to remain free of infection and therefore be vaccinated. The model estimates that with behavior change, 91% of individuals with the highest frequency of one-time sexual partnerships who were able/willing to be vaccinated were vaccinated, while without behavior change only 84% of these individuals were vaccinated. These estimates indicate that individuals with a high frequency of engaging in one-time partnerships were more likely to contract mpox virus before they had a chance to be vaccinated, and by delaying transmission, behavior change allowed for additional high-activity individuals to be vaccinated (Fig. S6).

### Infection Prevalence

Our analyses suggest that the outbreak will only end within a year with vaccination (Fig. 2B). After 1 year, we would have 77 [IQR 48 - 119] prevalent infections (defined as the number of infectious individuals on a given day) in the absence of any intervention. We estimated that vaccination would lower prevalent infections to 2 [IQR 0 - 4], or 0 [IQR 0 - 2] when paired with behavior change. Alternatively, behavior change alone would increase prevalent infection to 180 [IQR 116 - 292] one year into the outbreak. Behavior change can increase prevalence late in the outbreak because unlike vaccination, behavior change does not provide long-term protection. Vaccination replaces infection-driven herd immunity with vaccine-driven herd immunity, whereas behavior change only delays herd immunity. Thus, one year into the outbreak, more individuals are resistant to infection in the “no intervention” scenario” than in the “behavior change only” scenario, resulting in higher transmission, and thus infection prevalence. If vaccinations were not available, individuals may have reduced their sexual activity for longer periods of time than we observed. However, behavior change alone cannot end the outbreak within a year even when we extend the period of maximum behavior change by an additional two months (Fig. S7).

### Varying Vaccine Administration

The number and timing of vaccine allocations varied between jurisdictions, and there is uncertainty around vaccine efficacy. We estimated how changes in availability and timing could have impacted cases. We estimated that by one year into the outbreak, moving the initiation of vaccination earlier by 28 days would have decreased cumulative cases by 31% [IQR 46% - 13%], and moving the vaccination timeline later by 28 days would have increased cumulative cases by 29% [IQR 1% - 68%] (Fig. 3A). Increasing available vaccine doses by 50% would have decreased cumulative cases by 12% [IQR -23% - 33%], and decreasing doses by 50% would have increased cumulative cases by 52% [IQR 15% - 87%] (Fig. 3A). (Fig. 3B). Thus, for the parameter space explored, distributing vaccines earlier would linearly decrease cases, whereas increasing available vaccine doses would have diminishing returns.

**Figure 3:**
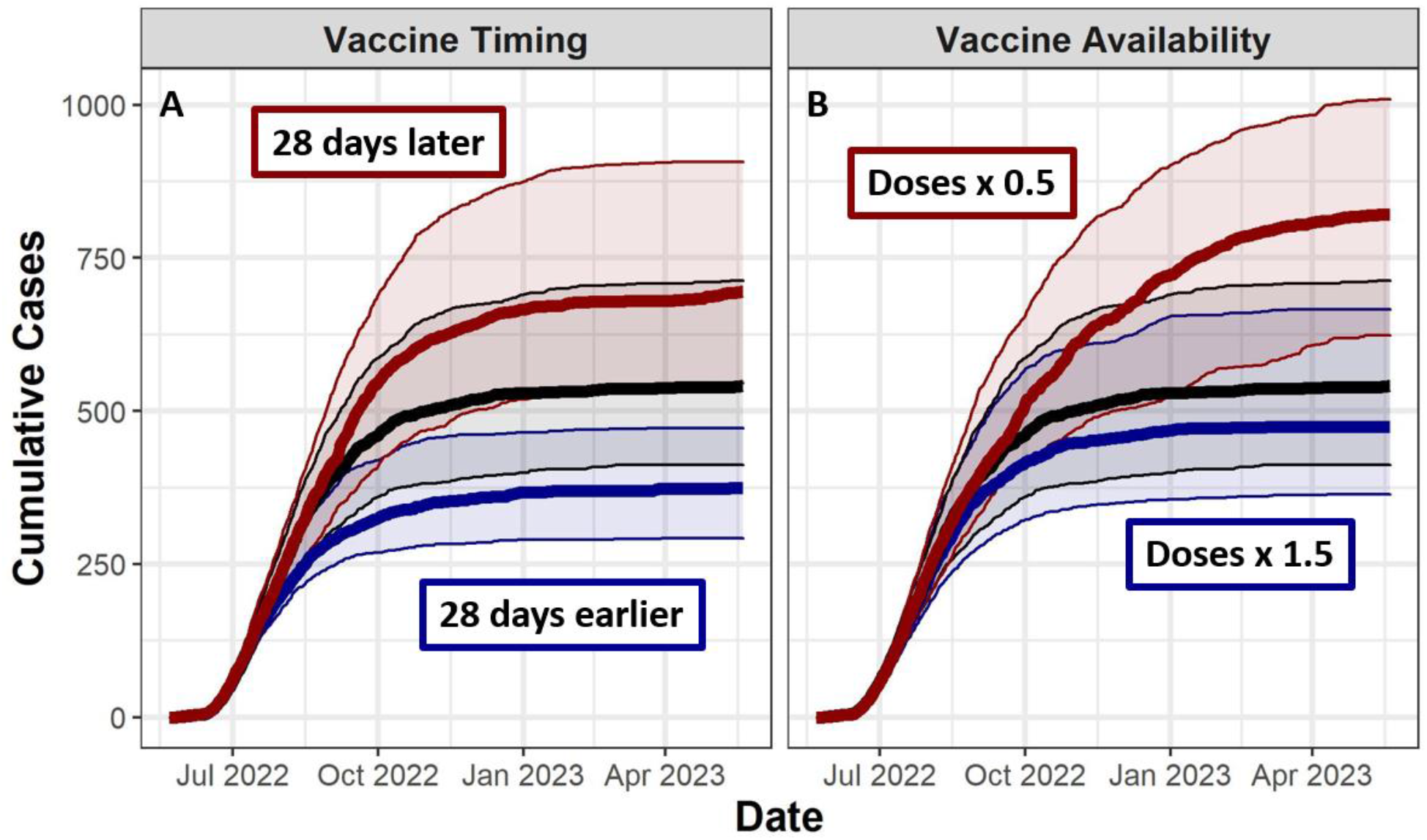
Changing number of administered doses and timing of vaccine administration can influence case reports. (A) Y-axis shows model estimates of cumulative case counts, over time on the X-axis from May 21^st^, 2022 to May 21^st^, 2023. Solid lines indicate median values from 120 simulations, while transparent bands outlined by thin lines represent interquartile ranges. Black lines show model results based on actual vaccine administration and a maximum vaccine efficacy of 90%, while red and blue lines represent more pessimistic and more optimistic scenarios, respectively. All scenarios shown here include behavior change.

## Discussion

We estimated the relative importance of vaccination and reductions in one-time sexual partnerships among MSM for averting mpox cases. We found that the majority of averted cases through October were averted by behavior change, but that vaccination began having an effect in September, was key to ending the outbreak in Washington D.C., and averted more cases overall. These results have several implications for the current mpox outbreak and for future outbreaks of sexually associated infections. First, national vaccine distribution is logistically time-consuming, and vaccines are not immediately effective. Thus, behavioral interventions can temporarily slow transmission, protecting communities from infection before vaccination takes effect. Partnering with affected communities is key to effective communication on behavioral interventions^19^.

During this outbreak, MSM, community-based organizations, digital and traditional media, and the public health community worked together in disseminating public health messaging and expanding vaccination^20–22^. The enthusiasm of MSM to protect themselves and the health of their communities^23^ highlights the effectiveness of public health efforts that manifest collaborations between affected communities and public health. Second, without vaccine administration, temporary behavioral interventions can reduce, but not eliminate, infection prevalence. This means that regions with low vaccine uptake might benefit from a focus on increasing vaccine administration even if cases are decreasing, and that behavior change alone is unlikely to successfully control future mpox outbreaks. Third, at early stages of an outbreak, administering vaccines sooner will provide substantial benefit in preventing cases over time and potentially have more impact than delayed administration of broader vaccination efforts that might lead to higher overall vaccine coverage.

Consistent with the current mpox outbreak, our model indicated that a combination of vaccination and behavior change would end the outbreak within a year. This may not be the case if (a) vaccines do not reach certain segments of the population, and (b) individuals who do not or cannot access vaccination preferentially partner with one another. We assumed in our model that only age and sexual positioning determines who individuals choose to partner with, and that neither of these factors influenced vaccination or treatment seeking. However, MSM partner with same-race individuals roughly 90% of the time^12^, and initial administration of the mpox vaccine was lower in Black individuals than in other racial groups^5^. Thus, mpox transmission might persist at a low but steady level in certain populations, mirroring our “behavior change only” scenario (Figure 2B), if vaccine equity is not improved.

Our results suggest that the outbreak ended in D.C. in large part due to vaccination and behavior change, rather than ending solely due to infection-driven herd immunity, shown to be a possibility in recent work^8^. Specifically, we find that without vaccination and behavior change, model fits that match the rapid early increase in cases overestimate the final number of cases, and model fits that match the final number of cases underestimate the initial growth of the epidemic. Possible differences in conclusions from these studies may arise from assumptions about the natural history of mpox (e.g. per-act transmission probability is assumed to be within the range of household transmission in^13^, much lower than our fitted parameter), or differences in the underlying sexual network.

### Limitations

The sexual network used here was primarily parameterized by surveys distributed in Atlanta in 2010^12^. The sexual networks of other cities may differ from that in Atlanta due to differences in demography, socioeconomic status, or cultural beliefs. Further, sexual networks in MSM may have changed in the last ten years due to increased prevalence of HIV pre-exposure prophylaxis and internet-based dating applications^24,25^. Thus, collecting new, regionally dispersed data on sexual networks of MSM and other populations experiencing high rates of sexually associated infections would improve modelling capabilities in future outbreaks. Another assumption we made is that sexual networks in D.C. are separate from neighboring jurisdictions. In reality, transmission in the network would be influenced by individuals traveling in and out of D.C., or by individuals who work in D.C. but live elsewhere.

Although substantial changes in sexual behavior by MSM in response to mpox have been documented^5^, we were unable to directly measure changes in one-time partnership behavior among MSM. Despite this, the initial decline in daily case reports was likely due to behavior changes among MSM because all parameter sets that fit the initial exponential increase in cases predicted that cases would have continued to sharply rise throughout the fall of 2022 unless we invoke behavior change. Our behavior analysis is further limited as we were only able to collect behavior proxies at the national, rather than regional level. However, it may be that MSM responded to national cases and news coverage, similar to how general interest in mpox (by Google search proxies) responded to national rather than local events. If the sexual activity levels of MSM respond to national rather than local events, then we expect that behavior change will have had a different impact on outbreaks in different cities, depending on the relative timing of the outbreak and national perception of mpox risk.

### Conclusions

This study indicates synergistic effects of sexual behavior change and vaccination in controlling the mpox outbreak. By changing their behavior gay, bisexual, and other men who have sex with men reduced transmission before vaccines were widely available, and those reductions in sexual behavior continue to protect individuals who are not vaccinated. We estimated that vaccination in turn will more effectively reduce infections later in the outbreak by providing long-term protection. The synergies observed when combining behavioral harm reduction and vaccination serve as a reminder for those administering mpox vaccination to use this clinical encounter to reinforce the importance of behavioral harm reduction strategies in a culturally appropriate way^4^.

## Data Availability

All data produced in the present study are available upon reasonable request to the authors

## Acknowledgements

The findings and conclusions in this report are those of the authors and do not necessarily represent the views of the Centers for Disease Control and Prevention or the National Cancer Institute.

## Author Contributions

Manuscript concept was designed by PAC, IHS, JM, YN. Model was designed by PAC, IHS, and EDP. JMA advised on model structure. NC, CEC, and KPD, provided input on modeling of behavior. DCP provided input on modeling of vaccination. WS and ATM provided epidemiological data from D.C. Manuscript was primarily written by PAC, with input from all authors.

## Citations

1. 2022 U.S. Map & Case Count | Monkeypox | Poxvirus | CDC. https://www.cdc.gov/poxvirus/monkeypox/response/2022/us-map.html.

2. Philpott, D. et al. Epidemiologic and Clinical Characteristics of Monkeypox Cases — United States, May 17–July 22, 2022. MMWR. Morb. Mortal. Wkly. Rep. 71, 1018–1022 (2022).

3. U.S. Monkeypox Case Trends Reported to CDC | Monkeypox | Poxvirus | CDC. https://www.cdc.gov/poxvirus/monkeypox/response/2022/mpx-trends.html.

4. Safer Sex, Social Gatherings, and Monkeypox | Monkeypox | Poxvirus | CDC. https://www.cdc.gov/poxvirus/monkeypox/prevention/sexual-health.html.

5. Delaney, K. P. et al. Strategies Adopted by Gay, Bisexual, and Other Men Who Have Sex with Men to Prevent Monkeypox virus Transmission — United States, August 2022. MMWR. Morb. Mortal. Wkly. Rep. 71, (2022).

6. Minhaj, F. S. et al. Monkeypox Outbreak — Nine States, May 2022. Morb. Mortal. Wkly. Rep. 71, 764 (2022).

7. Kriss, J. L. et al. Receipt of First and Second Doses of JYNNEOS Vaccine for Prevention of Monkeypox — United States, May 22–October 10, 2022. MMWR. Morb. Mortal. Wkly. Rep. 71, (2022).

8. Endo, A. et al. Heavy-tailed sexual contact networks and monkeypox epidemiology in the global outbreak, 2022. Science (80-.). 378, (2022).

9. Weiss, K. M. et al. Egocentric sexual networks of men who have sex with men in the United States: Results from the ARTnet study. Epidemics 30, 100386 (2020).

10. Jenness, S. M. et al. Impact of the Centers for Disease Control’s HIV Preexposure Prophylaxis Guidelines for Men Who Have Sex With Men in the United States. J. Infect. Dis. 214, 1800–1807 (2016).

11. Spicknall, I. H. et al. Modeling the impact of sexual networks in the transmission of Monkeypox virus among gay, bisexual, and other men who have sex with men — United States, 2022. vol. 71 (2022).

12. Hernández-Romieu, A. C. et al. Heterogeneity of HIV prevalence among the sexual networks of Black and White MSM in Atlanta: illuminating a mechanism for increased HIV risk for young Black MSM. Sex. Transm. Dis. 42, 505 (2015).

13. Beer, E. M. & Bhargavi Rao, V. A systematic review of the epidemiology of human monkeypox outbreaks and implications for outbreak strategy. PLoS Negl. Trop. Dis. 13, e0007791 (2019).

14. Grey, J. A. et al. Estimating the Population Sizes of Men Who Have Sex With Men in US States and Counties Using Data From the American Community Survey. JMIR Public Heal. Surveill 2016;2(1)e14 https://publichealth.jmir.org/2016/1/e14 2, pe5365 (2016).

15. FACT SHEET: Biden-Harris Administration’s Monkeypox Outbreak Response | The White House. https://www.whitehouse.gov/briefing-room/statements-releases/2022/06/28/fact-sheet-biden-harris-administrations-monkeypox-outbreak-response/.

16. Preliminary JYNNEOS Vaccine Effectiveness Estimates Against Medically Attended Mpox Disease in the U.S., August 15, 2022 – October 29, 2022 | Mpox| Poxvirus | CDC. https://www.cdc.gov/poxvirus/monkeypox/cases-data/mpx-JYENNOS-vaccine-effectiveness.html.

17. Sagy, Y. W. et al. Real-world effectiveness of a single dose of mpox vaccine in males. Nat. Med. 2023 1–1 (2023) doi:10.1038/s41591-023-02229-3.

18. Eaton, L. A. & Kalichman, S. C. Risk compensation in HIV prevention: Implications for vaccines, microbicides, and other biomedical HIV prevention technologies. Curr. HIV/AIDS Reports 2007 44 4, 165–172 (2007).

19. CDC’s Response to the 2022 Monkeypox Outbreak | Monkeypox | Poxvirus | CDC. https://www.cdc.gov/poxvirus/monkeypox/about/cdc-response.html.

20. Gonsalves, G. S., Mayer, K. & Beyrer, C. Déjà vu All Over Again? Emergent Monkeypox, Delayed Responses, and Stigmatized Populations. J. Urban Heal. 2022 994 99, 603–606 (2022).

21. Soelaeman, R. H. et al. Characteristics of JYNNEOS Vaccine Recipients Before and During a Large Multiday LGBTQIA+ Festival — Louisiana, August 9–September 5, 2022. MMWR. Morb. Mortal. Wkly. Rep. 71, 1379–1381 (2022).

22. Millman, A. J. et al. A Health Equity Approach for Implementation of JYNNEOS Vaccination at Large, Community-Based LGBTQIA+ Events — Georgia, August 27– September 5, 2022. MMWR. Morb. Mortal. Wkly. Rep. 71, 1382–1883 (2022).

23. McNaghten, A. D. et al. COVID-19 Vaccination Coverage and Vaccine Confidence by Sexual Orientation and Gender Identity — United States, August 29–October 30, 2021. Morb. Mortal. Wkly. Rep. 71, 171 (2022).

24. Paz-Bailey, G. et al. Trends in Internet Use Among Men Who Have Sex With Men in the United States. J. Acquir. Immune Defic. Syndr. 75, S288 (2017).

25. Wu, H. et al. Uptake of HIV Preexposure Prophylaxis Among Commercially Insured Persons—United States, 2010–2014. Clin. Infect. Dis. 64, 144–149 (2017).

26. Loosier, P. S. et al. Reddit on PrEP: Posts About Pre-exposure Prophylaxis for HIV from Reddit Users, 2014–2019. AIDS Behav. 26, 1084–1094 (2022).

