## appendix for "Modelling the impact of vaccination and sexual behavior change on reported cases of mpox in Washington D.C"

Parameterizing Behavior Change

We chose to analyze activity in individual subreddits (individual conversation boards on reddit, a social media discussion based website) as a proxy for risk awareness following prior work on HIV awareness[26]. We first searched for LGBTQ+ focused subreddits that (a) had more than 10,000 members, and (b) contained at least 5 posts or comment threads containing "monkeypox", "mpox", or "mpx". We identified 7 sites: r/askgaybros, r/askgaybrosover30, r/askgaymen, r/gay, r/gaybros, r/lgbt, r/lgbtnews. For each of these subreddits, we collected posts by date, and the number of comments on each post, filtering out early posts concerning first reports of mpox in non-U.S. countries (Fig. S1A). A day that includes many posts with many comments indicates a high level of interest in mpox among the observed LGBTQ+ community. However, not all comments on a post occur on the day that the post is made, particularly since posts receiving many comments will stay on the "front page" of a subreddit for several days, meaning that they will be among the first post that site visitors see. Thus, to represent the interest over time that a single post can represent, we fit an exponential decay curve to the number of comments received each day by the most commented post containing "monkeypox" posted on these subreddits. We then spread the number of comments made on each post over days according to this exponential decay (Fig. S1B). Because posts that attract comments are shown to more users, the number of comments a post receives may be a exponential function of interest in a topic. Thus, we take a log transform of comments per day (Fig. S1C). Finally, we take a 2 week running average of log(comments) to generate risk perception among MSM over time (Fig. S1D). The maximum relative reduction in probability of one-time partnerships per day in our model (40%) occurs on the day of maximum reddit activity (July 30th). This relative decrease is then scaled over time with reddit activity, e.g. we model a 20% reduction in one-time partnership probability on July 11th and September 24th, when reddit activity was half of the maximum value.

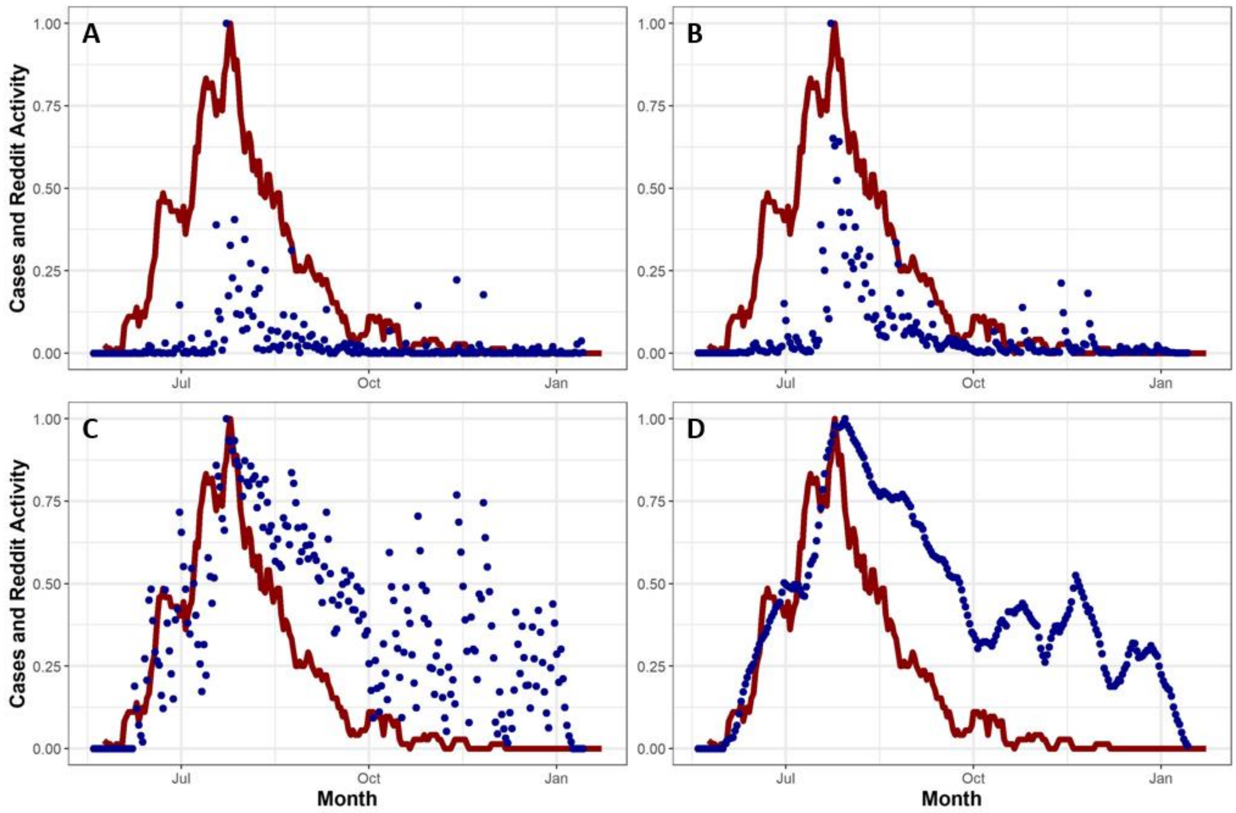


**Figure S1:** Here, we transform reddit data into our model's relative risk perception metric. For the red line in all panels, the Y-axis represents 1 week running average of case reports from Washington D.C., set relative to 1, over months on the X-axis. For blue dots, Y-axis represents (A) number of posts + number of comments in examined subreddits containing mpox related terms, relative to 1, (B) Number of posts and comments, taking into account the exponential decay of comments over time, (C) the log transform of panel B, (D) the running 2 week average of panel C.

These the subreddits are not specific to D.C., and thus we needed to quantify regional variation in online mpox interest to ascertain whether subreddit searches needed to be corrected for regional interest. Thus, we examined the proportion of Google searches containing "monkeypox", "mpox", or "mpx" from cities which experienced early mpox outbreaks (D.C., New York, and Chicago) compared to cities that experienced later outbreaks or that did not experience a substantial outbreak (Atlanta, Houston, Phoenix, Seattle). We find that Google searches for these terms generally do not vary by location within the U.S. (Fig. S2), indicating that the timing of the general population's risk perception of mpox did not depend on local trends. Further, we find that Google search interest peaks at a similar point to reddit activity. Thus we conclude that mpox risk perception is synchronous nationally, meaning that national trends can be used as a proxy for local risk perception.

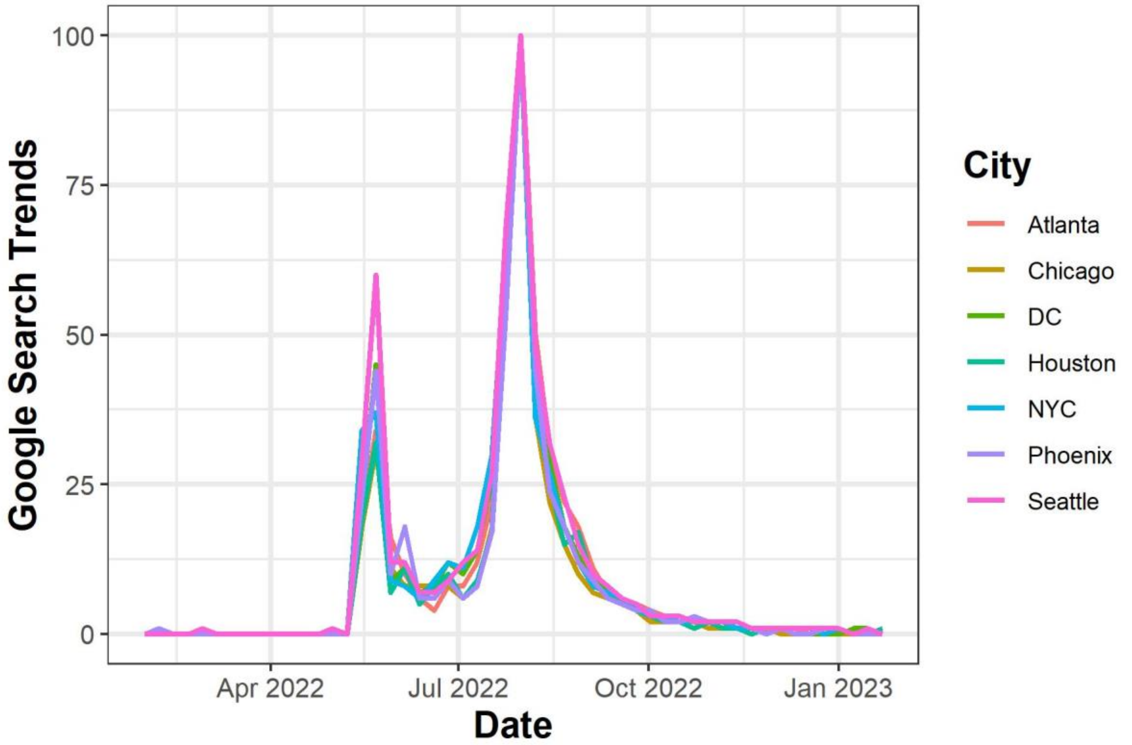


**Figure S2:** Google search trends for "monkeypox" do not vary by location. The Y-axis represents the proportion of all Google searches from a location which contained the term "monkeypox", relative to 1, over time on the X-axis. Colors indicate search location. The early peak corresponds to early news reports on international mpox cases and initial cases reported in the United States. The main peak corresponds with the declaration of mpox as a public health emergency.

Fitting procedure

We drew most model parameters from published literature (Table S1). However, we fit five unknown parameters to Washington D.C. case data: the probability of transmission per contact ($\mu$), the length in days of the pre-symptomatic infectious period ($l$), the number of individuals infected with mpox virus at the Capital Pride Parade/Festival, occurring on June 11-12 ($\varepsilon$), the maximum percent reduction in probability of one-time sexual contact per day in response to the mpox outbreak ($\omega$), and the proportion of administered vaccine doses reported by Washington D.C. that went to the modeled population rather than non-msm or non-resident commuters. When fitting to D.C. case data, we removed all case reports occurring in cisgendered women (defined as case reports where both sex at birth and gender identity were reported as female).

To increase the efficiency of our fitting procedure, we fit parameters over two different time periods. We first ran the model from May 21 – Jul 10. This period encompasses the Capital Pride Parade/Festival and the period during which individuals who contracted mpox at pride would develop symptoms and report for testing. However, this period occurs before large numbers of individuals were vaccinated or a large proportion of the population were aware of the risk of mpox (by our reddit post metric). Thus, we set behavior change and vaccination to zero, and fit all other parameters to this 50 day period. We conducted a grid search, running the model across a range of values for these three parameters ($\mu$=[0.6,0.90], $l$=[1,4], $\varepsilon$=[0,60]). We then selected the combination of parameters that minimized the negative log likelihood (nll) of median cumulative cases over time in the model generating observed cumulative cases over time reported in Washington D.C. (using a Poisson distribution). When fitting, we ran each parameter combination for 60 simulations. We performed this procedure to a parameter interval of 0.1 for $\mu$, 10 for $\varepsilon$, and 1 for $l$. We found that over this time period the best fit parameter set included an 80% probability of transmission per sex act, a 4 day latent period, and 40 individuals infected at D.C. pride.

We thus set $\varepsilon = 40$, $\mu = 0.8$ and $l = 4$ and then ran our model from May 21 – December 10. This time period includes the decline of daily case reports, and so should inform intervention parameters. Thus, we again conducted a grid search, running the model across a range of values for maximum behavior change and proportion of reported vaccines administered to the modeled population ($\omega$=[0.3,0.7], $\theta$=[0.5,1.0]). We performed this procedure to a parameter interval of 0.1 for $\mu$ and $\theta$. We found that the best fitting parameter set slightly underestimated case reports in the early stages of the outbreak, and slightly overestimated case reports in the later stages of the outbreak. Thus, we varied $\mu$ and $\omega$ over smaller intervals, before maximizing likelihood at $\varepsilon = 40$, $\mu = 0.875$, $l = 4$, $\omega = 0.4$ and $\theta = 1.0$.

To test the likelihood of the outbreak ending entirely due to infection-driven herd immunity, we repeated the above fitting process while holding behavior change and vaccine administration at 0 ($\omega = 0$ and $\theta = 0$).

Table S1: model parameters

| **Parameter Description** | **Value** | **Source** |
|---|---|---|
| Initial Conditions | | |
| Population size | 37,400 | Grey et al. 2016 |
| Initial number infectious individuals | 5 | Model Assumption |
| Date seeded with initial infections | May 21th | Case report data |
| Number individuals infected at D.C. pride | 40 | Fit to case report data |
| Sexual Network Parameters | | |
| Proportion individuals with 0 main and 0 casual partners | 0.471 | Jenness et al. 2017 |
| Proportion individuals with 0 main and 1 casual partners | 0.167 | Jenness et al. 2017 |
| Proportion individuals with 0 main and 2 casual partners | 0.074 | Jenness et al. 2017 |
| Proportion individuals with 1 main and 0 casual partners | 0.220 | Jenness et al. 2017 |
| Proportion individuals with 1 main and 1 casual partners | 0.047 | Jenness et al. 2017 |
| Proportion individuals with 1 main and 2 casual partners | 0.021 | Jenness et al. 2017 |
| Mean duration of Main partnerships | 407 days | Jenness et al. 2017 |
| Mean duration of Casual partnerships | 166 days | Jenness et al. 2017 |
| Probability per day of individual with 0 casual and 0 main partners having a one-time sexual contact | 0.0093 | Jenness et al. 2017, Weiss et al. 2020 |
| Probability per day of individual with 1 casual and 0 main partners having a one-time sexual contact | 0.012 | Jenness et al. 2017, Weiss et al. 2020 |
| Probability per day of individual with 2 casual and 0 main partners having a one-time sexual contact | 0.012 | Jenness et al. 2017, Weiss et al. 2020 |
| Probability per day of individual with 0 casual and 1 main partners having a one-time sexual contact | 0.0080 | Jenness et al. 2017, Weiss et al. 2020 |
| Probability per day of individual with 1 casual and 1 | 0.0079 | Jenness et al. 2017, Weiss et al. 2020 |

| | | |
|---|---|---|
| main partners having a one-time sexual contact | | |
| Probability per day of individual with 2 casual and 1 main partners having a one-time sexual contact | 0.0079 | Jenness et al. 2017, Weiss et al. 2020 |
| Probability per day of individual in activity group 1 having a one-time sexual contact | 0 | Jenness et al. 2017 |
| Probability per day of individual in activity group 2 having a one-time sexual contact | 0.001 | Jenness et al. 2017 |
| Probability per day of individual in activity group 3 having a one-time sexual contact | 0.0054 | Jenness et al. 2017 |
| Probability per day of individual in activity group 4 having a one-time sexual contact | 0.01 | Jenness et al. 2017 |
| Probability per day of individual in activity group 5 having a one-time sexual contact | 0.032 | Jenness et al. 2017 |
| Probability per day of individual in activity group 6 having a one-time sexual contact | 0.29 | Weiss et al. 2020 |
| Proportion of the population in activity groups 1-5 each | 0.19 | Jenness et al. 2017, Weiss et al. 2020 |
| Proportion of the population in activity group 6 | 0.05 | Jenness et al. 2017, Weiss et al. 2020 |
| Mean square root of age difference between Main partners | 0.464 | Jenness et al. 2017 |
| Mean square root of age difference between Casual partners | 0.586 | Jenness et al. 2017 |
| Mean square root of age difference between one-time partners | 0.544 | Jenness et al. 2017 |
| Proportion of population who are exclusively insertive | 0.242 | Jenness et al. 2017 |

| | | |
|---|---|---|
| Proportion of population who are exclusively receptive | 0.321 | Jenness et al. 2017 |
| Proportion of population who are versatile | 0.437 | Jenness et al. 2017 |
| Probability of having sexual contact per timestep with Main partner | 0.22 | Jenness et al. 2017 |
| Probability of having sexual contact per timestep with Casual partner | 0.14 | Jenness et al. 2017 |
| Probability of having sexual contact per timestep with one-time partner | 1 | Jenness et al. 2017 |
| Natural History Parameters | | |
| Probability of transmission per sexual contact | 0.875 | Fit to case report data |
| Mean duration of post-exposure, pre-symptomatic period | 7.6 days | Charniga et al. 2022 |
| Mean duration of pre-symptomatic infectious period | 4 days | Fit to case report data |
| Mean duration of infectious period | 27 days | Spicknall et al. 2022 |
| Treatment Seeking and Vaccine Parameters | | |
| Probability of seeking treatment | 0.8 | Farley et al. 2003 |
| Duration of infectious period if seeking treatment (time until medical attention post-symptoms) | 15 days on day 1, linearly decreases to 5.0 days on day 42, stays at 5.0 days for rest of simulation. | Figure S3 |
| Vaccine efficacy before second dose | 37% | Payne et al. 2022 |
| Vaccine efficacy after second dose | 69% | Payne et al. 2022 |
| Minimum Time between first and second dose | 28 days | |
| Behavior Change Parameters | | |
| Maximum percent reduction in daily probability of forming one-time sexual partnerships. | 40% | Fit to case report data |

Table S2: Vaccination first-dose administration data for Washington D.C. Doses in first four rows contain only doses used for post-exposure prophylaxis and so were not included in our model.

| Start Date | End Date | First Doses Given | Second Doses Given |
|---|---|---|---|
| May 29, 2022 | June 4, 2022 | 1 | 0 |
| June 5, 2022 | June 11, 2022 | 23 | 0 |
| June 12, 2022 | June 18, 2022 | 18 | 0 |
| June 19, 2022 | June 25, 2022 | 16 | 0 |
| June 26, 2022 | July 2, 2022 | 319 | 0 |
| July 3, 2022 | July 9, 2022 | 627 | 15 |
| July 10, 2022 | July 16, 2022 | 938 | 13 |
| July 17, 2022 | July 23, 2022 | 3179 | 32 |
| July 24, 2022 | July 30, 2022 | 3433 | 64 |
| July 31, 2022 | August 6, 2022 | 4481 | 78 |
| August 7, 2022 | August 13, 2022 | 2723 | 279 |
| August 14, 2022 | August 20, 2022 | 1303 | 377 |
| August 21, 2022 | August 27, 2022 | 1739 | 2791 |
| August 28, 2022 | September 3, 2022 | 1060 | 3674 |
| September 4, 2022 | September 10, 2022 | 994 | 2485 |
| September 11, 2022 | September 17, 2022 | 924 | 2050 |
| September 18, 2022 | September 24, 2022 | 486 | 2049 |
| September 25, 2022 | October 1, 2022 | 165 | 325 |
| October 2, 2022 | October 8, 2022 | 129 | 388 |
| October 9, 2022 | October 15, 2022 | 101 | 335 |
| October 16, 2022 | October 22, 2022 | 83 | 235 |
| October 23, 2022 | October 29, 2022 | 82 | 155 |
| October 30, 2022 | November 5, 2022 | 69 | 139 |
| November 6, 2022 | November 12, 2022 | 51 | 87 |
| November 13, 2022 | November 19, 2022 | 39 | 77 |
| November 20, 2022 | November 26, 2022 | 22 | 44 |
| November 27, 2022 | December 3, 2022 | 30 | 70 |
| December 4, 2022 | December 10, 2022 | 29 | 61 |

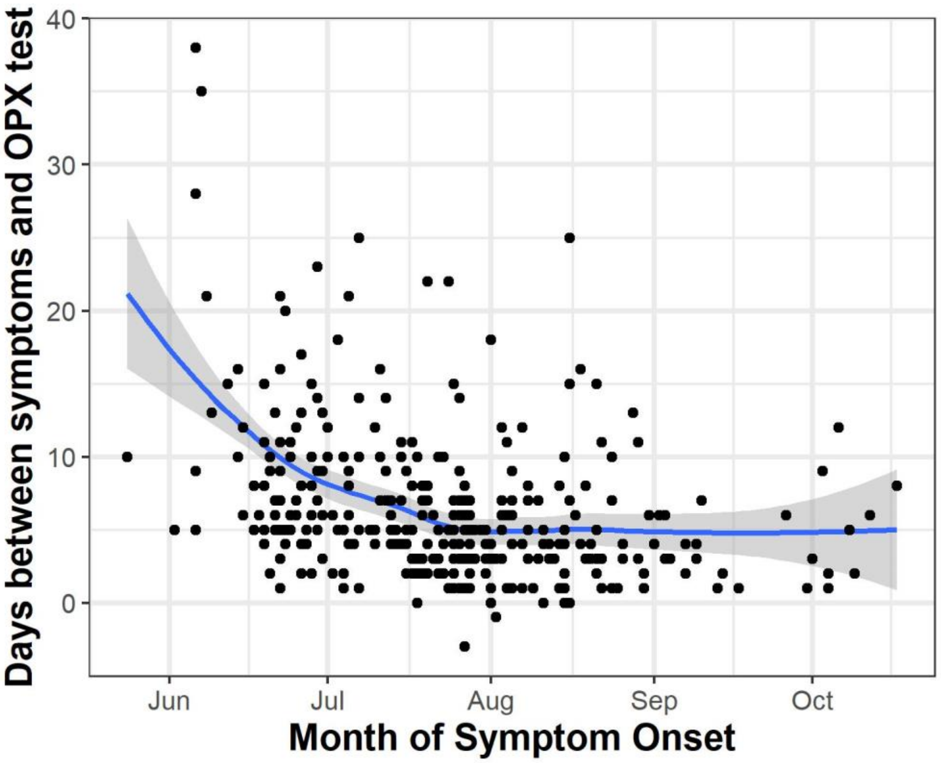


**Figure S3:** Time between symptom onset and being tested for orthopoxvirus (OPX) decreased over the course of the outbreak. Y-axis shows the delay between symptom onset and testing for orthopoxvirus, over month of illness onset on the X-axis from May 24$^{th}$, 2022 to Oct. 17$^{th}$, 2022. Points represent individual case reports, while the blue line and transparent band represent mean and 95% confidence interval of the smoothed function of the mean. All case reports are from Washington D.C.

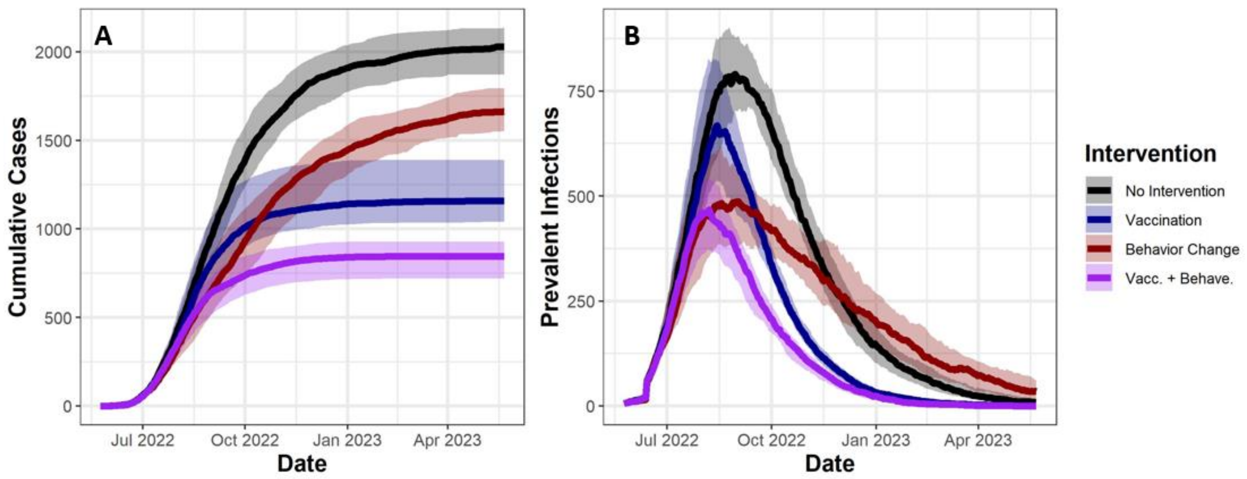


**Figure S4:** Here, we shrink the effective population size in our model from 37,400 to 20,000 as we assume that a proportion of MSM are not involved in the relevant sexual mixing pool. This means that vaccinations can reach a larger portion of the population, and that infection-driven herd immunity will be achieved faster. We find that the our core finding are unchanged: behavior change averts cases before vaccination, but vaccination averts more cases overall. (A) Y-axis shows model estimates of cumulative cases (i.e. individuals who are diagnosed with mpox), over time on the X-axis from May 21st 2022 to May 21st 2023. (B) Y-axis shows prevalent infections, over time on the X-axis. Solid lines indicate median values from 120 simulations, while transparent bands represent interquartile ranges. Colors indicate intervention combinations.

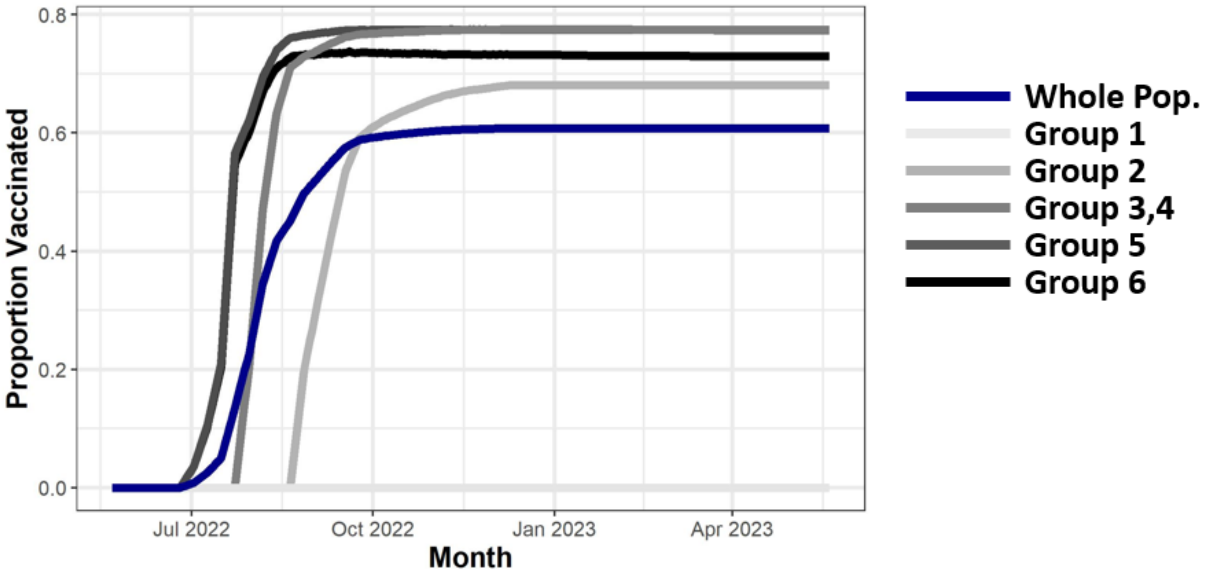


**Figure S5:** Proportion of sexual activity groups vaccinated over time. Y-axis shows proportion of the whole population or individual activity groups (group 1 = lowest activity, group 6 = highest activity) that are vaccinated with either 1 or 2 doses, over time on the X-axis. Groups 3 and 4 share 1 line because their trends overlap.

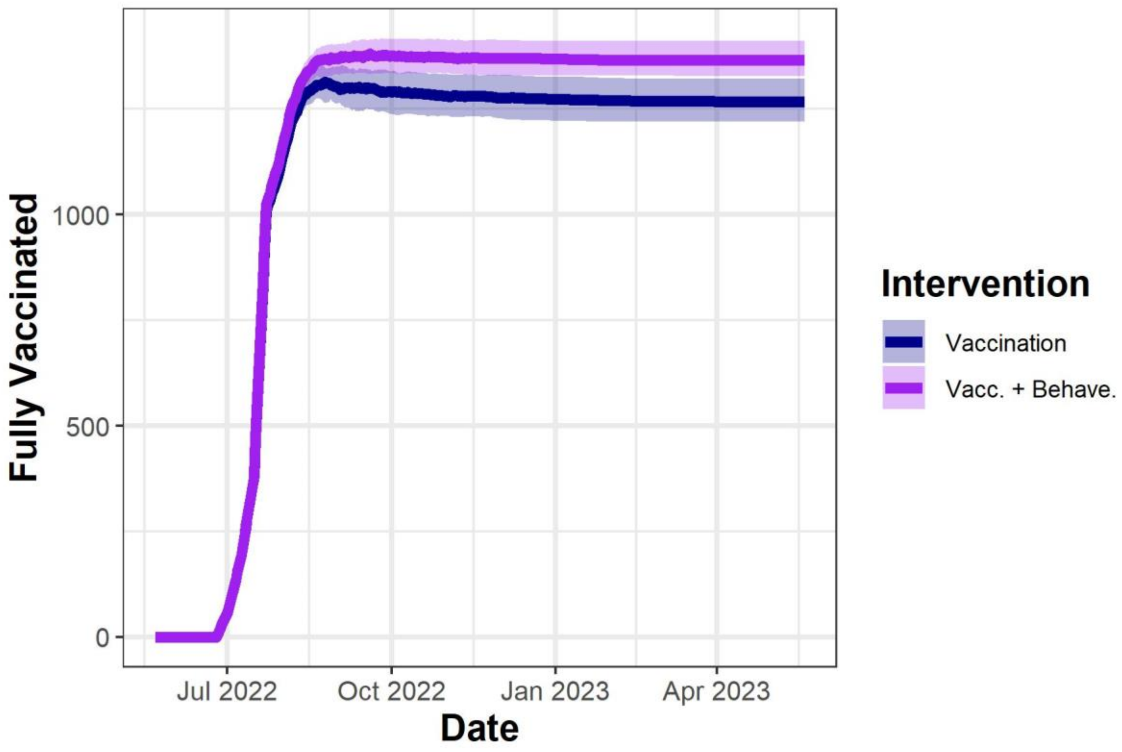


**Figure S6:** Y-axis shows the number of individuals in the most sexually active group who are fully vaccinated, over date on the X-axis. Solid lines indicate median values from 120 simulations, while transparent bands outlined by thin lines represent interquartile ranges. Decreases in number of individuals who are fully vaccinated occurring from August 2022 onwards represent individuals who experience break-through infections, and thus are no longer counted in the vaccinated class.

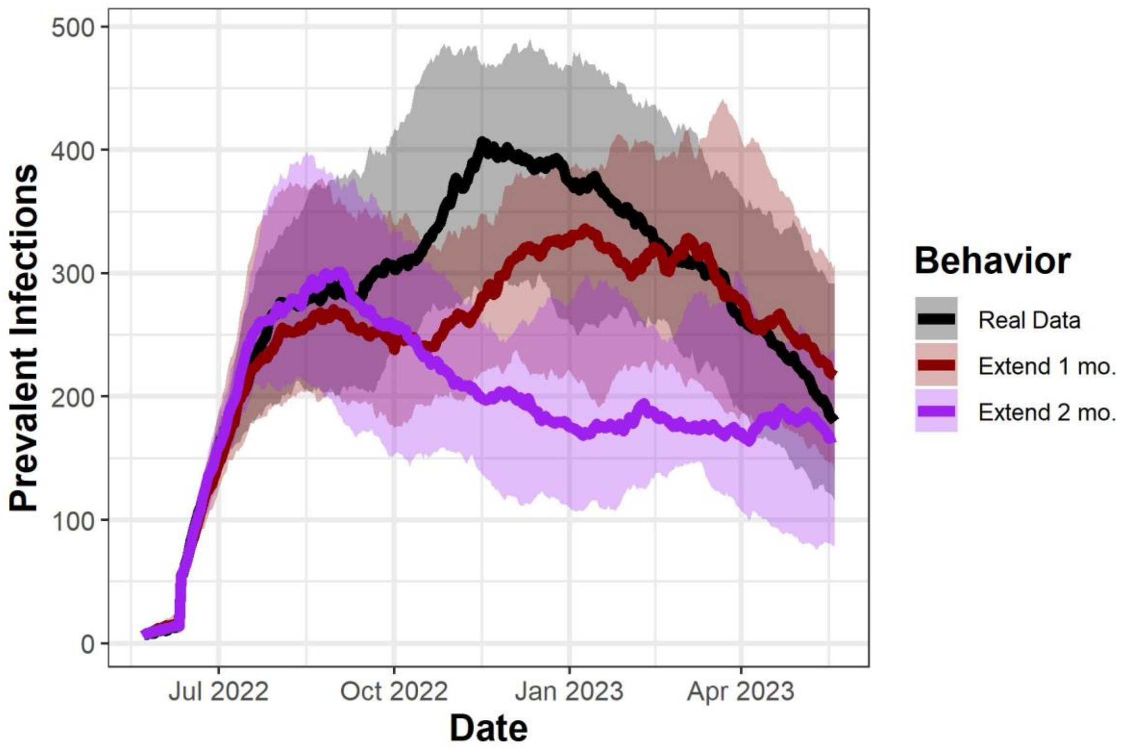


**Figure S7:** Behavior change alone would not have been able to end the outbreak within a year, even when we extend the time period of maximim reduction of one-time partnership probabilities. Y-axis shows prevalent infections, over time on the X-axis. Solid lines indicate median values from 120 simulations, while transparent bands represent interquartile ranges. Colors indicate the length of time over which individuals maintain a 40% reduction in one-time partnership probabilities. Black indicates data based on reddit activity, for red we extend maximum partnership reduction by one month, and for purple we extend maximum partnership reduction by two months.

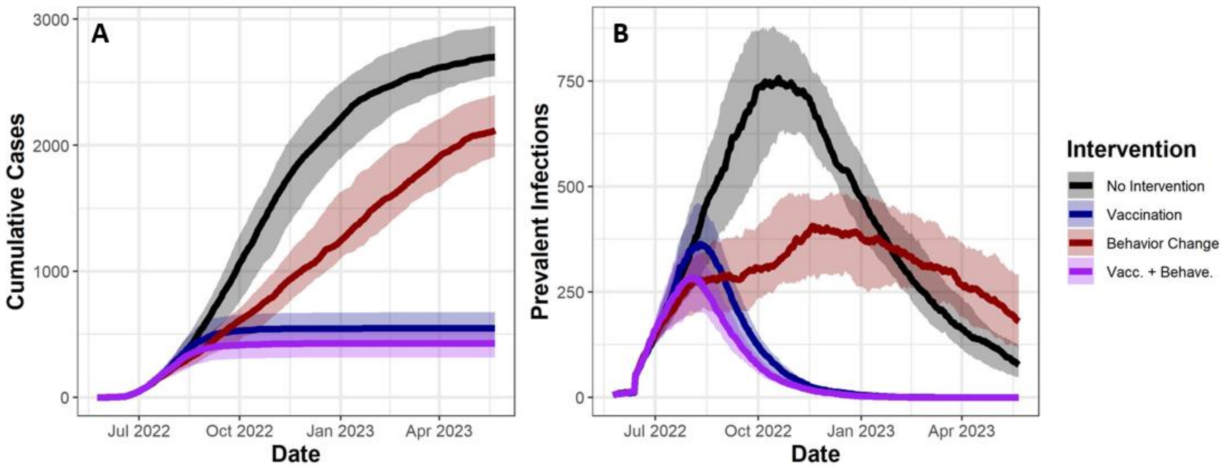


**Figure S7:** Here, we assume a vaccine efficacy of 85% after first dose and 95% after second dose based on [17], rather than 37% and 69% vaccine effectiveness, respectively, used in the main text [16]. We find that the our core finding are unchanged: behavior change averts cases before vaccination, but vaccination averts more cases overall. (A) Y-axis shows model estimates of cumulative cases (i.e. individuals who are diagnosed with mpox), over time on the X-axis from May 21st 2022 to May 21st 2023. (B) Y-axis shows prevalent infections, over time on the X-axis. Solid lines indicate median values from 120 simulations, while transparent bands represent interquartile ranges. Colors indicate intervention combinations.
